# Communicating polygenic and non-genetic risk for atherosclerotic cardiovascular disease - An observational follow-up study

**DOI:** 10.1101/2020.09.18.20197137

**Authors:** Elisabeth Widén, Nella Junna, Sanni Ruotsalainen, Ida Surakka, Nina Mars, Pietari Ripatti, Juulia J Partanen, Johanna Aro, Pekka Mustonen, Tiinamaija Tuomi, Aarno Palotie, Veikko Salomaa, Jaakko Kaprio, Jukka Partanen, Kristina Hotakainen, Pasi Pöllänen, Samuli Ripatti

## Abstract

**Background:** Algorithms including both traditional risk factors and polygenic risk scores (PRS) can significantly improve prediction of atherosclerotic cardiovascular disease (ASCVD). However, the clinical benefit of adding PRS to clinical risk evaluation remains unclear.

**Objectives:** The study evaluated the attitudes of 7,342 individuals (64% women, mean age 56 yrs) upon receiving personal genome-enhanced ASCVD risk information, and prospectively assessed the impact on the participants’ health behavior.

**Methods:** The participant’s 10-year risk for ASCVD was estimated using both a traditional clinical risk score and a PRS-enhanced score, and both scores were communicated directly to study participants with an interactive web-tool.

**Results:** When reassessed after 1.5 years by a clinical visit and questionnaires, 20.8% of individuals at high (>10%) ASCVD risk had seen a doctor, 12.4% reported weight loss, 14.2% of smokers had quit smoking, and 15.4% had signed up for health coaching online. Altogether, 42.6% of individuals at high risk had made some health behavioral change compared to 33.5% of persons at low/average risk such that a higher baseline risk predicted a favorable change (p<0.001), with both clinical (p<0.001) and genomic factors (p=0.003) contributing independently. Seeing a doctor and weight loss both resulted in clinically significant improvement of lipid profiles (lower LDL-cholesterol and triglycerides) and lower systolic blood pressure (p<0.01).

**Conclusions:** Web-based communication of personal ASCVD risk-data including polygenic risk to middle-aged persons can motivate positive changes in health behavior. It supports integration of genomic information into clinical risk calculators as a feasible approach to enhance disease prevention.

**Condensed Abstract:** Prediction tools that combine polygenic risk scores (PRS) with clinical factors provide a new opportunity for improved risk assessment and prevention of atherosclerotic cardiovascular disease (ASCVD), but the clinical impact of PRS has hitherto remained unclear. We evaluated the longitudinal effects of using a web-based tool to communicate genome-based ASCVD risk-information to 7,342 middle-aged individuals. 42% of persons at high risk improved their health behavior during follow-up which resulted in clinically significant improvement of lipid profiles and lower systolic blood pressure. This supports integration of PRS into clinical risk calculators as a feasible approach to enhance disease prevention.

## Introduction

Atherosclerotic cardiovascular disease (ASCVD) is the leading cause of premature death and disability world-wide, the majority of which is attributed to coronary heart disease (CHD) and stroke ^1^. Hitherto prevention strategies have relied on two related approaches, one being to decrease the average risk factor levels in the population, which has been highly successful in reducing overall premature morbidity and mortality from ASCVD in the past 50 years^2^. This approach essentially ignores interindividual differences, of which some arise from biology, some from social stratification, and therefore does not address inequalities in health very efficiently.

The other approach focuses on managing modifiable risk factors in persons at high overall risk, and clinical guidelines for disease prevention recommend personal risk assessment or the use of quantitative risk scores utilizing well-established risk markers, e.g. blood pressure and cholesterol levels, to aid in multifactorial risk estimation ^3^. Nonetheless, a large proportion of cases is erroneously classified at low or intermediate risk by current risk prediction methods ^4^, and this has led to continuous efforts to identify new predictive biomarkers to enhance accuracy of the prediction.

While few significant markers have emerged, numerous novel genetic loci impacting the risk for CHD, and polygenic risk scores (PRS) have been shown to significantly predict the disease risk, and to capture risk information that is orthogonal to traditional risk factors ^5,6^. Thus, prediction tools that combine genomic information with clinical factors provide a new opportunity for improved disease risk assessment and prevention^7^. So far, genome-wide information has not been utilized routinely for ASCVD-prediction in the clinical setting. In particular, it remains unknown how knowing one’s personal genomic risk for ASCVD may impact on the health behavior and propensity to seek care.

Thus, in order to enhance clinical disease prediction and to empower individuals to improve their health behavior, we developed a web-based tool for personal ASCVD risk-assessment, based on both genomic and traditional risk factors, and tested the longitudinal impact of using the tool to communicate disease risk information directly to 7,342 individuals. The goals of the study were: 1) to develop a web-based tool for communicating the genomic and clinical risks for participants and test its feasibility in a large cohort of middle-aged persons, 2) to assess the study participants’ attitudes upon directly receiving personal disease risk information, and 3) to examine the impact of receiving genome-based disease risk-information on health behavior.

## Methods

### The GeneRISK study-cohort

The GeneRISK-study is a prospective observational study including 7,342 45-65-year old individuals from Southern Finland. The recruitment was carried out during 2015 – 2017 at three different sites. From the Kymenlaakso province in South-Eastern Finland (CAREA - Kymenlaakso social and health care services) we randomly identified 4,857 individuals from the population register and invited them by mail. In addition, 1,369 customers of Mehiläinen Oy, a private provider of health care and occupational health care (Helsinki and Turku region offices), and 1,116 blood donors (Finnish Red Cross Blood Service in Helsinki) were recruited through online advertising directed to the Mehiläinen and Finnish Red Cross Blood Service customers respectively. Individuals with previous history of ASCVD, pregnant women, or individuals under guardianship were excluded from the study.

After consenting to the study, participants underwent a clinical health check-up. Information regarding their disease history, smoking habits, family history of CHD, and marital status, and education (Supplemental information) was collected via electronic questionnaires. Plasma glucose and serum lipids were determined from overnight fasting samples, and the genetic profile was analyzed using the HumanCoreExome BeadChip (Illumina Inc, San Diego, CA, USA). Genotypes were called using GenomeStudio and zCall at the Institute for Molecular Medicine Finland (FIMM), phased using SHAPEIT2, and imputed using IMPUTE2 and a combined reference panel of 1000 Genomes Phase I integrated haplotypes and 1,943 Finnish genomes (https://www.biorxiv.org/content/10.1101/080770v1).

Two estimates of the personal 10-year risk for ASCVD were generated for each participant, 1) based on clinical risk factors only (age, sex, smoking status, total cholesterol (TC), HDL-cholesterol (HDL-C), systolic blood pressure (SBP), current usage of antihypertensive medication and family history of early-onset CHD) and 2) based on the combined effects of the clinical risk factors and a SNP-based CHD-risk polygenic score encompassing roughly 49,000 markers ^6,8,9^. Following Tikkanen et al, ASCVD was defined as myocardial infarction, unstable angina pectoris, coronary revascularization (coronary artery bypass graft or percutaneous transluminal coronary angioplasty), death due to CHD, or ischemic stroke events. The CHD PRS was estimated as the sum of the risk alleles weighted by the logarithms of the odds-ratio estimates taken from the CardiogramPlusC4D meta-analysis summary statistics^6^. The ASCVD-risk estimates were interpreted as outlined in the Finnish National Guidelines for prevention of coronary heart disease and/or stroke, i.e. a 10-year risk <2% is considered small, a risk between 2 and <10% is average, a risk between 10 and <15% is high, and a risk of ≥15% is considered very high.

The study participants were given access to their personal disease risk information through a web-based personal health record (PHR), which was specifically set up for this research project. Once the ASCVD-risk prediction was completed, the study participants were sent a text message instructing them to log on to the PHR where they could view their ASCVD-risk results with KardioKompassi®, an interactive web-tool developed in house, and all their laboratory values. In case the participant’s ASVCD was considered high (10-year risk ≥10%) KardioKompassi advised the person to see his/her personal doctor to discuss the results. Participants with 10-year risk <10% were advised to maintain a healthy lifestyle. In the PHR, participants were further given access to an online library service, maintained by The Finnish Medical Society Duodecim, which provided general information on medical conditions and information on how to obtain treatment and care. Moreover, all participants were offered the opportunity to sign up for online health coaching services also offered by The Finnish Medical Society Duodecim free of charge. The online services included coaching modules supporting smoking cessation, weight control, healthy dietary habits, exercise, stress reduction, and healthy sleeping.

On average one and a half year (16.9±5.7 months) after receiving their personal results the study participants were invited to a follow-up visit, including a health check and fasting plasma and lipid measurements. At follow-up, study participants also answered e-questionnaires assessing their attitudes towards receiving personal disease risk information, and their health behavior. Positive health behavior at follow-up was primarily assessed based on the questionnaire, e.g. seeing a doctor to discuss the personal ASCVD risk results and smoking cessation. The outcome of the doctor’s visit, e.g. initiation of blood lipid lowering medication or blood pressure lowering medication, was also assessed by the questionnaire. Weight change during follow-up was assessed both using the questionnaire and the measured weight at the baseline and follow-up visits. Individuals who both had reported weight loss and whose measured weight was at least 0.1 kg lower at follow-up compared with their weight at baseline were considered as having lost weight. Signing up for health coaching was defined using activity log information of the PHR.

The GeneRISK study was carried out according to the principles of the Helsinki declaration and the Council of Europe’s (COE) Convention of Human Rights and Biomedicine. All study participants gave their informed consent to participate in the study, to receive data on their personal ASCVD-risk, including genomic risk information, and to be re-contacted. Participants’ DNA, blood, serum, and plasma samples, in addition to their demographic information and health data have been stored in the THL Biobank (https://www.thl.fi/en/web/thlfi-en/topics/information-packages/thl-biobank). The study protocol was approved by the Ethical Committee of the Helsinki and Uusimaa Hospital district.

### Statistical analyses

The differences of the health behavior changes (signing up for health coaching online, weight loss, quitting smoking, seeing physician or any of the above) between high and low risk individuals were tested using **χ**^2^-test.

Logistic regression was used to estimate the effects of polygenic and clinical risk for ASCVD to positive health behavior in the same models during follow-up. Two models were performed, one by adjusting for age and sex only and another including also marital status, education, having viewed the personal ASCVD –risk results in KardioKompassi at least once, and clinical center in the model.

Changes in blood lipids, plasma glucose and blood pressure during follow up in individuals at high 10-year risk for ASCVD both according to whether study participants had discussed their ASCVD-risk results with their physician or not, and whether study had a weight loss during follow-up or not were tested using multiple linear regression. These models were adjusted for age, sex, marital status, education, having opened KardioKompassi at least once, clinical center, and the baseline level of the corresponding risk factor.

Statistical analyses were done using the R Version 3.6.2 (2019-12-12).

## Results

### Baseline risk assessment

Altogether 2,651 men and 4,691 women participated in the health checkup at baseline. Their clinical and behavioral characteristics are shown in Table 1. Majority of participants were overweight or obese (37.7% of women and 48.2% of men were overweight (BMI 25-30 kg/m^2^), and 24.4 vs. 24.4% obese (BMI ≥ 30 kg/m^2^)), 69.1% had total cholesterol exceeding the recommended level (> 5 mmol/l). Of all individuals, 11.1% received lipid lowering therapy whereas 22.0% were treated for hypertension.

**Table 1.** Clinical characteristics of study participants at baseline.

|  | Men | Women |
| --- | --- | --- |
| N | 2,651 | 4,691 |
| Age (years) | 55.9 ± 5.9 | 55.7 ± 5.7 |
| BMI (kg/m<sup>2</sup>) | 27.7 ± 4.4 | 27.2 ± 5.3 |
| BMI ≥ 30 (N (%)) | 646 (24.4%) | 1143 (24.4%) |
| Waist circumference (cm) | 100.6 ± 12.6 | 90.7 ± 13.4 |
| Total cholesterol (mmol/l) | 5.5 ± 1.0 | 5.6 ± 1.0 |
| HDL-C* (mmol/l) | 1.43 ± 0.45 | 1.79 ± 0.50 |
| LDL-C† (mmol/l) | 3.4 ± 0.9 | 3.3 ± 0.9 |
| S-triglycerides (mmol/l) | 1.49 ± 1.18 | 1.15 ± 0.65 |
| ApoA1 (mmol/l) | 1.58 ± 0.26 | 1.75 ± 0.30 |
| ApoB (mmol/l) | 1.06 ± 0.27 | 0.97 ± 0.26 |
| Systolic blood pressure (mmHg) | 134.6 ± 15.6 | 126.6 ± 16.7 |
| Diastolic blood pressure (mmHg) | 88.3 ± 10.0 | 84.3 ± 9.9 |
| Daily smokers (N (%)) | 328 (12.4%) | 451 (9.6%) |
| Lipid-lowering medication (N (%)) | 380 (14.3%) | 437 (9.3%) |
| Blood pressure-lowering medication (N (%)) | 642 (24.2%) | 972 (20.7%) |

Data are presented as mean ± SD
\*HDL-C = high-density lipoprotein cholesterol
†LDL-C = low-density lipoprotein cholesterol

The distribution of the participants’ 10-year risk for ACVD-risk is shown in Figure 1. Applying the risk thresholds outlined by the Finnish national guidelines for disease prevention, 24.0% of study participants were at high risk for developing ACVD, i.e. a 10-year absolute risk exceeding 10%. More specifically, 7.7% had a 10-year risk between 10 and 15%, 5.4% a 10-year risk of 15% or more. In accordance with the Finnish national disease prevention guidelines, individuals with previously diagnosed diabetes, anamnesis of angina pectoris or heart failure, fasting glucose > 7 mmol/l, or serum LDL-cholesterol exceeding 6 mmol/l were automatically considered to be at high risk, and in such a case KardioKompassi advised the person to discuss the ASCVD risk with his/her doctor without separately computing an individual 10-year ASCVD risk estimate. Altogether 10.9% of study participants were in this category. Of the remaining individuals at high risk, 40.7% were smokers, 17.0% were on lipid lowering therapy, and 30.2% were obese with a BMI >30 kg/m^2^.

**Figure 1.**
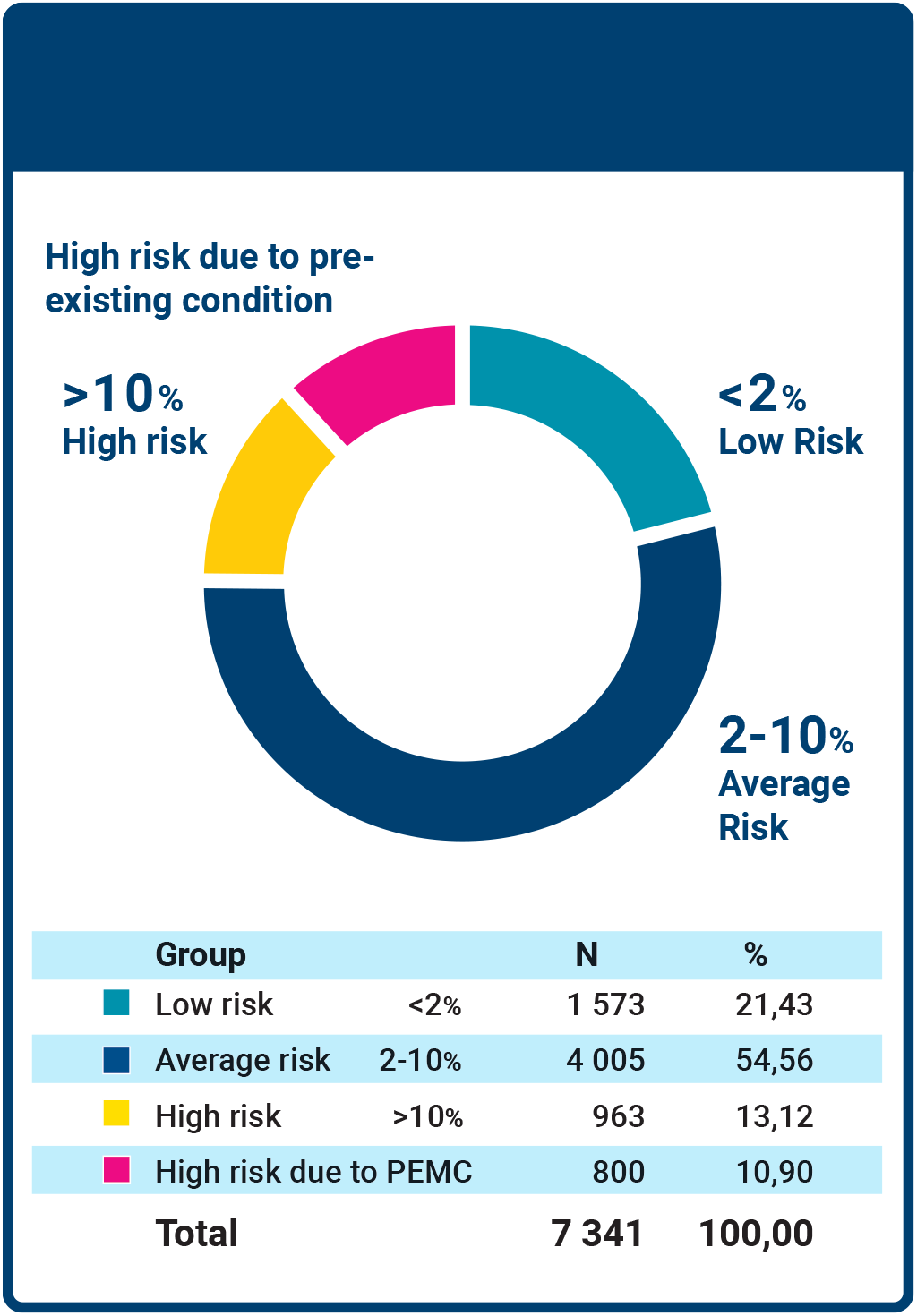
Distribution of the 10-year ASCVD-risk across the GeneRISK –cohort at baseline. PEMC = Pre-existing medical condition impacting on the risk for atherosclerotic cardiovascular disease, e.g. diabetes, heart failure, angina pectoris, severe hyperlipidemia (LDL-cholesterol > 6 mmol/l)

### Disease risk communication

The personal ASCVD-risk information was communicated and interpreted to all study participants by KardioKompassi®, a web-application developed in-house, which presents the personal disease risk data both as a point estimate of the absolute disease risk and as a function of age (Figure 2). It also includes an interactive component allowing the user to explore how changes of modifiable risk factors may impact on the overall disease risk (more detailed information in the Supplement). Analyses of the participants’ usage of the web service showed that 89.7% (n=6,586) chose to view their KardioKompassi data at least once. Sex, marital status and education level all associated with the likelihood of viewing the results, i.e. women (p<0.001), persons married or in a relationship (p=0.0077), or persons with higher education (p<0.001) more often retrieved their results than others (Supplementary Table 3).

**Figure 2.**
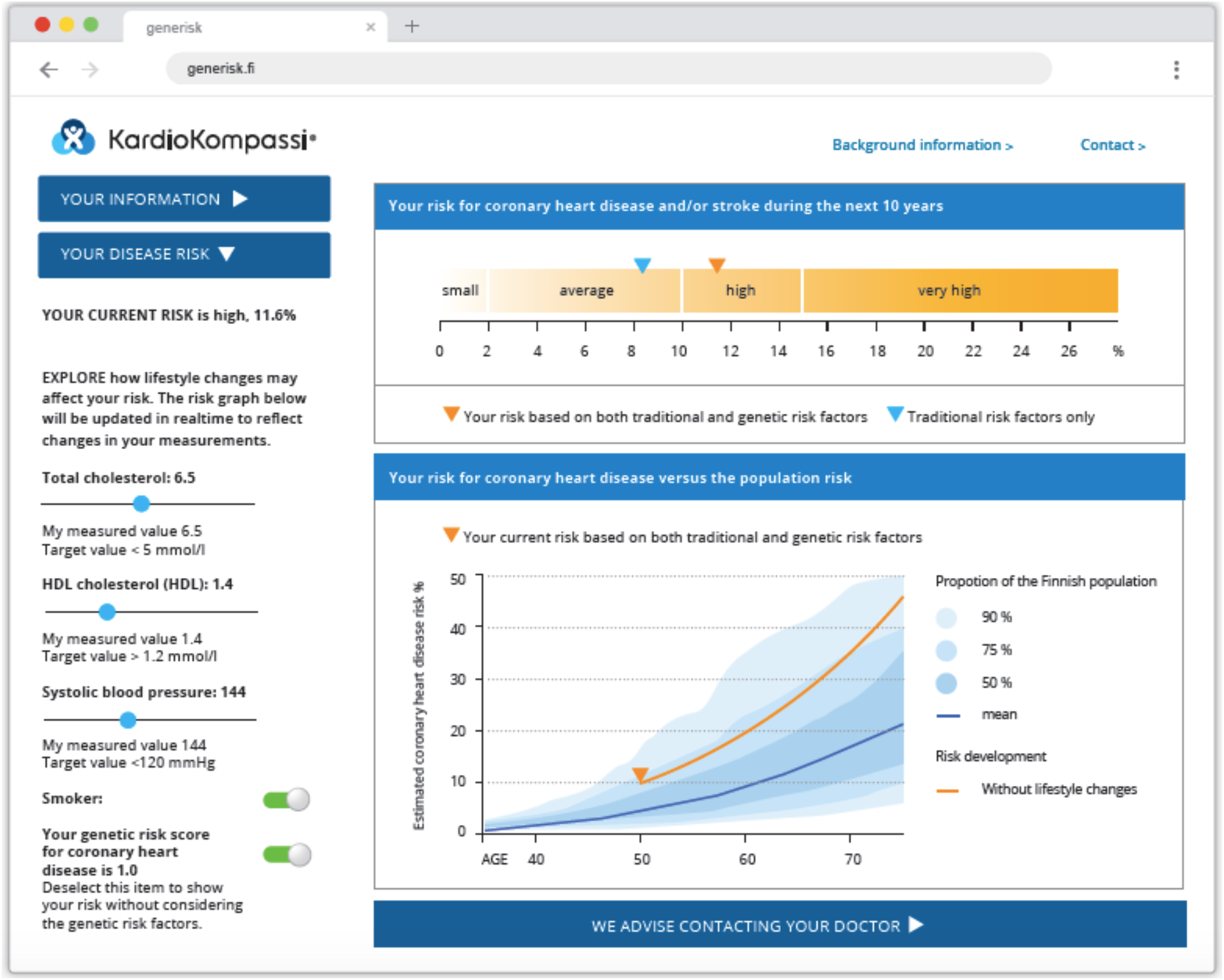
A screenshot from the KardioKompassi® risk prediction application. Figure 2A. The figure shows a detail of the GUI displaying a male individual’s risk. The upper panel shows the overall risk for coronary heart disease and/or stroke and the lower panel shows the risk for CHD as a function of age (y-axis). The x-axis in the lower panel shows the estimated risk for CHD, the solid orange line indicates the joint risk including both traditional and genomic risk factors, the blue background shows the CHD-risk in the Finnish reference population (the blue line indicates the mean population risk).

**Figure 2B.**
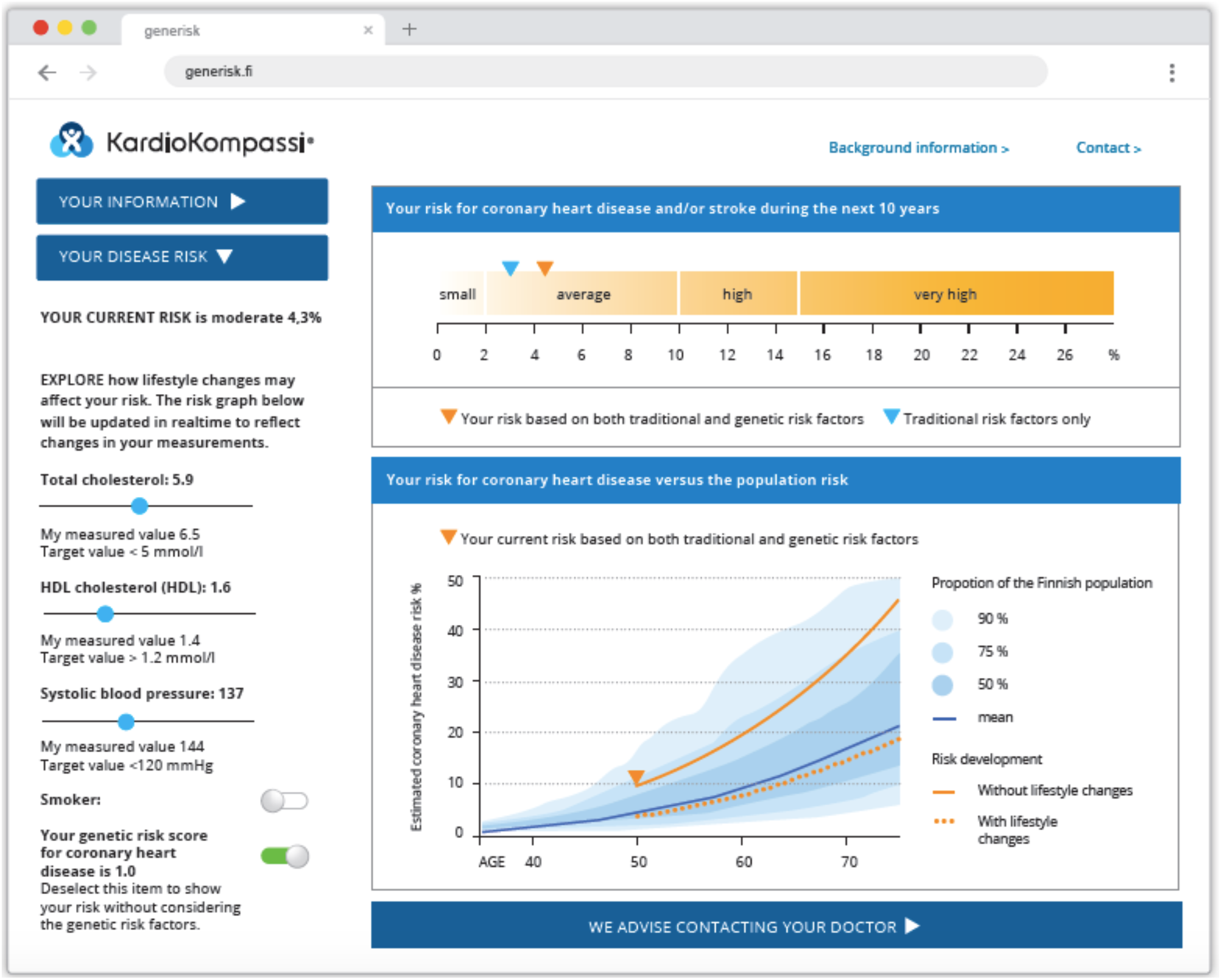
The figure shows interactive feature of the KardioKompassi-application, and depicts the results of the same individual as in Figure 2A. Using the sliders displayed on the left-hand side of the KardioKompassi GUI, the participants could test how a change of the modifiable risk factors influences on his/her the overall risk. The broken orange line shows the disease risk provided that the study participant doesn’t smoke, the total cholesterol was 0.5 mmol/l lower, the HDL-cholesterol was 0.2 mmol higher, and the systolic blood pressure was 7 mmHg lower.

### Assessment at the follow-up visit

On average 17 months (16.9±5.7 months) after having received their personal KardioKompassi-report we invited study participants to a second health checkup and asked them to fill out an e-questionnaire probing their health behavior and attitudes. Seventy-one percent (n=5,191) of individuals seen at baseline participated in the follow-up study. When asked about their attitude towards receiving personal disease risk information, almost 90% said that the information was easy to understand and that the results were useful (Table 2). Twenty-two percent indicated that their results had been unexpected, whereas a third reported that they had received concerning results. As many as 97% stated that they believe that their ASCVD-risk is influenced significantly by genetic factors. Nonetheless, this belief did not discourage them from undertaking actions to improve their health. On the contrary, 99% of participants thought that they can impact on their risk through lifestyle choices. Moreover, 89% indicated that their personal risk information motivates them to take better care of their health. Seventy-five percent of the participants also believed that physicians know how to interpret and utilize genomic information in their clinical practice.

**Table 2.** Study participants’ attitudes concerning their personal disease risk information assessed at 18 months of follow-up (n=5,191).

|  | All | Men | Women |
| --- | --- | --- | --- |
| My personal disease risk information was easy to understand | 88% | 87% | 88% |
| I received useful information regarding my personal disease risk | 90% | 90% | 90% |
| I received unexpected disease risk results | 22% | 23% | 21% |
| I received concerning disease risk results | 29% | 29% | 29% |
| I believe that genetic factors importantly influence my disease risk | 97% | 96% | 97% |
| I believe that I can impact on my disease risk through my lifestyle | 99% | 99% | 99% |
| My personal risk information motivates me to take better care of my health | 89% | 89% | 90% |
| Clinical doctors know how to interpret and utilize genome information | 75% | 79% | 72% |

### Changes in health behavior and risk factors during follow-up

Making significant or lasting changes in health behavior is known to be challenging ^10-12^. Therefore, we next examined the changes in health behavior the study participants had undertaken during follow-up. At baseline, everyone was offered access to online health coaching services and persons at elevated ASCVD-risk had also been advised to see a doctor. Given that a substantial proportion of persons at elevated risk were obese or smokers, we selected the four following outcomes to indicate positive health behavior change: 1) signing up for health coaching online, 2) weight loss, 3) smoking cessation, and 4) seeing a physician during follow-up. We then characterized the individuals at high risk (10-year ASCVD risk >10%) and compared their health behavior with individuals at average or low risk (<10%; Table 3).

**Table 3.** Comparison of health behavior during follow-up between individuals at high risk for atherosclerotic cardiovascular disease (ASCVD-risk ≥ 10%) and persons at average or low risk for atherosclerotic cardiovascular disease (ASCVD-risk <10%).

| | ASCVD-risk $\geq 10\%$<br>n = 683 | ASCVD-risk $<10\%$<br>n = 3,993 | P-value |
| --- | --- | --- | --- |
| Seen a physician (%) | 20.8 | 8.3 | P < 0.001 |
| Weight loss<br>(% of study participants) | 12.4 | 8.67 | 0.002 |
| Quit smoking<br>(% of smokers) | 14.2 | 10.9 | 0.3 |
| Signed up for health<br>coaching online | 15.4 | 20.2 | 0.004 |
| **Any of the above (%)** | **42.6** | **33.5** | **P < 0.001** |

The data showed that 15.4% of individuals at high risk had signed up for health coaching online, 12.4% reported weight loss (mean (SD) weight loss −3.9 kg (± 2.8) kg), 14.2% of smokers had quit smoking by their own self-report and 20.8% had seen a doctor (Table 3). While individuals at average or low risk more often signed up for health coaching than persons at high risk, (20.2 vs 15.4%, p=0.004), a higher proportion of persons at high risk reported weight loss or saw a physician. Taken together, 42.6% of individuals at high risk for ASCVD had taken some action to lower their disease risk compared to 33.5% of individuals in the average/low risk-group (p<0.001) (Table 3). A higher ASCVD risk at baseline was associated with a positive change in health behavior during follow-up, to which both a higher clinical and genomic risk score contributed independently (Table 4). The data further showed that the improvement in health behavior also resulted in improved clinical risk factor profiles. Individuals at high ASCVD-risk who either saw a physician or lost weight both showed significant improvement of blood lipids and blood pressure (Table 5 and 6).

**Table 4.** The independent contribution of polygenic risk for coronary heart disease (CHD) and absolute clinical risk for ASCVD to positive health behavior during follow-up Response variable: Positive health behavior*

| Explanatory variable | Adjusted | OR (CI) | P-value |
| --- | --- | --- | --- |
| Polygenic risk for CHD | Age, sex, clinical risk | 1.10 (1.03-1.17)<sup>†</sup> | **0.004** |
|  | Multivariate | 1.10 (1.03-1.17)<sup>‡</sup> | **0.003** |
| Clinical risk for ASCVD | Age, sex, polygenic risk | 1.42 (1.28-1.59)<sup>§</sup> | **P < 0.001** |
|  | Multivariate | 1.53 (1.37-1.72)<sup>?</sup> | **P < 0.001** |

\* Positive health behavior includes at least one of the following: signing up for health coaching online, weight loss, smoking cessation, or visiting a doctor, yes = 1,625, no = 3,046.
<sup>†</sup> Adjusted for age, sex, and the absolute clinical risk for ASCVD
<sup>‡</sup> Multivariate: adjusted for age, sex, the absolute clinical risk for ASCVD, clinic clinical center, whether opened KardioKompassi at least once, marital status, and education level.
<sup>§</sup> Adjusted for age, sex, and polygenic risk for CHD.
<sup>?</sup> Multivariate: adjusted for age, sex, polygenic risk for CHD, clinical center, whether opened KardioKompassi at least once, marital status and education level.

**Table 5.** Changes in blood lipids, blood glucose and blood pressure during follow up in individuals at elevated 10-year risk for ASCVD according to whether study participants had discussed their ASCVD-risk results with a doctor.

| **Risk factor** | **Visited a doctor** | **Baseline (mean(sd))** | **Follow-up (mean(sd))** | **Difference (mean(sd))** | **p-value\*** |
| --- | --- | --- | --- | --- | --- |
| Total cholesterol (mmol/l) | Yes | 5.9 (0.9) | 5.3 (1.1) | -0.6 (1.3) | 0.006 |
|  | No | 5.7 (1.0) | 5.5 (1.0) | -0.2 (1.0) |  |
| LDL-cholesterol (mmol/l) | Yes | 3.9 (0.9) | 3.3 (1.0) | -0.5 (1.1) | 0.009 |
|  | No | 3.6 (0.8) | 3.4 (0.9) | -0.2 (0.8) |  |
| HDL-cholesterol (mmol/l) | Yes | 1.35 (0.37) | 1.40 (0.38) | 0.052 (0.24) | 0.95 |
|  | No | 1.30 (0.37) | 1.35 (0.40) | 0.057 (0.25) |  |
| S-Triglycerides (mmol/l) | Yes | 1.6 (0.9) | 1.4 (0.7) | -0.2 (0.7) | 0.041 |
|  | No | 1.7 (0.9) | 1.6 (0.8) | -0.1 (0.7) |  |
| Apolipoprotein A (mmol/l) | Yes | 1.6 (0.3) | 1.6 (0.3) | -0.0 (0.2) | 0.29 |
|  | No | 1.5 (0.2) | 1.5 (0.3) | -0.0 (0.2) |  |
| Apolipoprotein B (mmol/l) | Yes | 1.2 (0.2) | 1.0 (0.3) | -0.1 (0.3) | 0.001 |
|  | No | 1.1 (0.3) | 1.1 (0.3) | -0.0 (0.2) |  |
| Systolic blood pressure (mmHg) | Yes | 143.5 (17.3) | 135.9 (17.9) | -6.5 (15.1) | 0.007 |
|  | No | 139.4 (15.8) | 137.5 (15.4) | -2.2 (13.8) |  |
| Diastolic blood pressure (mmHg) | Yes | 92.1 (11.3) | 88.8 (10.3) | -3.0 (8.5) | 0.09 |
|  | No | 90.3 (9.3) | 89.1 (9.4) | -1.3 (8.4) |  |
| Blood glucose (mmol/l) | Yes | 5.8 (0.4) | 5.9 (0.6) | 0.04 (0.5) | 0.94 |
|  | No | 5.8 (0.5) | 5.9 (0.6) | 0.06 (0.5) |  |

Yes, n=143; No, n=542
\*p-values are adjusted for the risk factor level at baseline

**Table 6.**
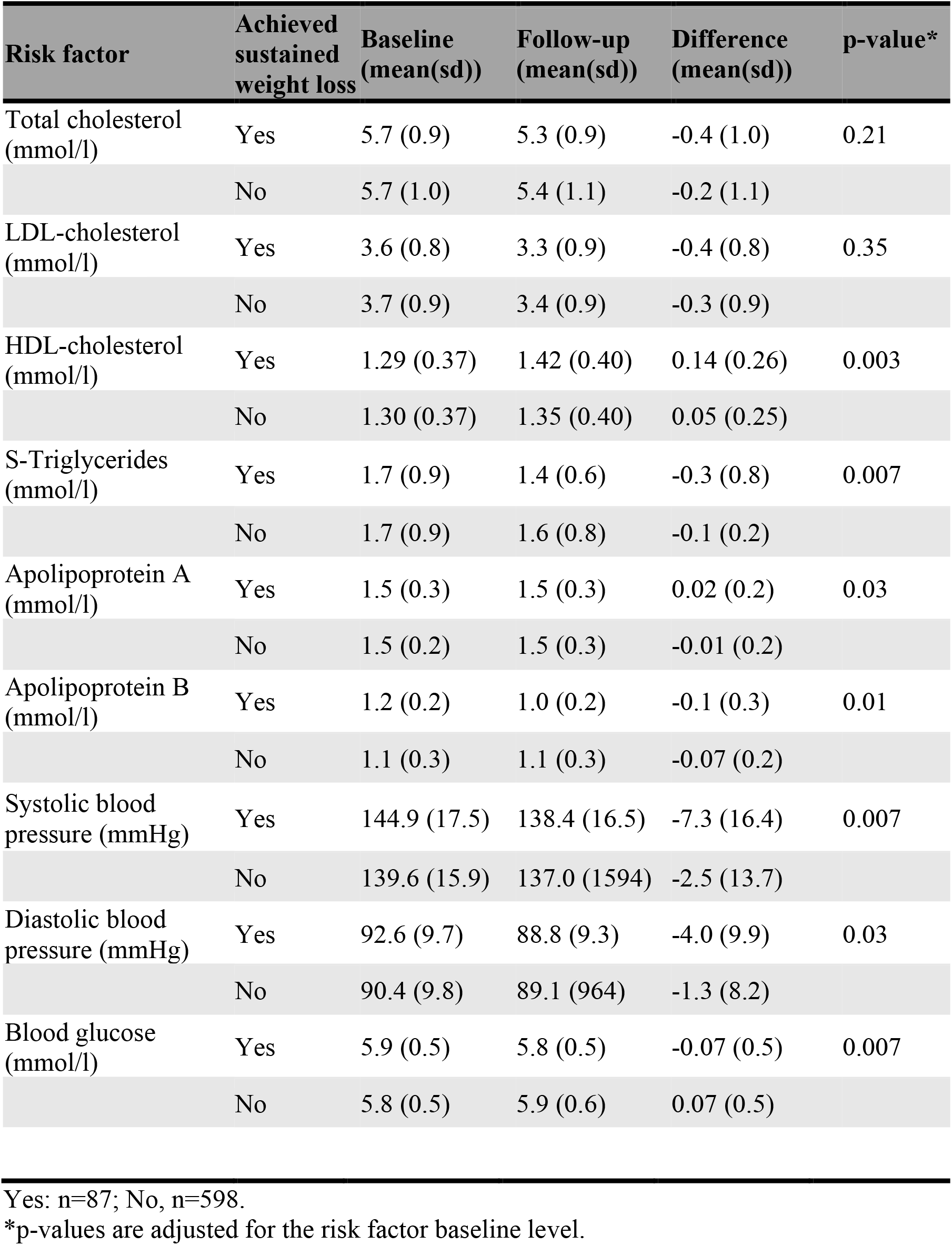
Changes in blood lipids, blood glucose and blood pressure during follow up in individuals at elevated 10-year risk for ASCVD according to whether study participants reported weight loss during follow-up

Based on the e-questionnaire information, 47.7% of high-risk individuals who had discussed their results with their physician had been recommended follow-up of lipid levels, 24.6% had been prescribed lipid-lowering medication, 50.7% had been recommended follow-up of blood pressure levels, and 23.1% had been prescribed blood pressure lowering medication. As a consequence, among high risk individuals, physician visits were associated with a 0.5 mmol/l reduction of LDL-cholesterol, and a 6.5 and 3.0 mmHg reduction of systolic and diastolic blood pressure, respectively (Table 5). Also, individuals who reported weight loss showed improvement of their lipid and blood pressure profiles, i.e. on average a 0.14 mmol/l increase in HDL-cholesterol, 0.3 mmol/l reduction of serum triglycerides, and 7.3 and 4.0 mmHg reduction of systolic and diastolic blood pressure respectively (Table 6). Although individuals who quit smoking gained weight on average (1.9 ± 4.4 kg), their lipid or blood pressure levels did not change significantly during follow-up.

## Discussion

Here we present results from the largest prospective study to date assessing changes in health behavior in individuals who received personal information on their genome-based risk for ASCVD. The study participants accessed their data with a novel interactive web-based tool and reported 1) that their risk results were easy to understand, 2) that they believe that both their genome and their behavioral choices impact on their disease risk, and 3) that the personal risk information motivates them to take better care of their health. The study also shows that 42% of individuals at high risk for ASCVD undertook action to reduce their disease risk, and that elevation of both clinical and genomic risk factors at baseline were independently associated with the positive change of health behavior during follow-up.

### The clinical role of PRS:s

Despite the growing pool of evidence supporting that polygenic risk information significantly improves the accuracy of ASCVD risk prediction, the clinical impact of the PRS has remained unclear. Earlier studies focusing on single or a small number of common genetic variants with low effect size suggest that disclosure of genetic information alone has little impact on desired health behavior, such as changing diet, increasing physical activity, or smoking cessation^13^. However, the clinical use of genetic risk information for primary prevention will likely require the incorporation of a PRS into existing screening protocols and prevention strategies that jointly consider the multiple factors impacting on the disease risk. Therefore, we chose to build the population-based GeneRISK-study on national prevention guidelines for ASCVD-prevention. The results provide us with compelling evidence supporting that the tested approach is both feasible and effective.

### Feasibility and attitudes

First, an overwhelming majority of study participants found their personal disease risk information both useful and easy to understand. One out of three participants also reported that they were concerned about their results, but as expected, concerning or unexpected results were most frequently reported by individuals at elevated risk for ASCVD (Supplementary Table 5). Overall, the participants seemed to have recognized the multifactorial nature of the disease given that the vast majority considered both genomic factors and lifestyle factors as important contributors to their personal risk. This finding is in accordance with a previous study which evaluated the relationship between belief in the genetic etiology of heart disease and cancer and awareness of lifestyle causes. That study reported that persons who believe that genetics influence the disease risk, in fact, are more likely to believe that also lifestyle plays a role ^14^.

### Overall impact on health behavior

Secondly, more than two-fifths of individuals at high risk for ASCVD in the current study took action to reduce their risk during follow-up. The higher the risk at baseline the more likely a change in health behavior during follow-up, but most importantly, both the information on clinical and the genomic risk factors independently contributed to this behavioral change. Smaller-sized studies, including a few hundred participants, have previously reported that disclosure of PRS for coronary heart disease associates with increased perception of personal control ^15^, in addition to increased information seeking and sharing ^16^. In accordance with these findings our study further adds that receiving personal genome-based disease risk information motivates individuals at elevated disease risk more than others, and show that a substantial proportion of individuals at high risk also undertake relevant actions to promote their health. However, our results are in contrast with a previous study of a selected sample of blood donors, where disclosing genetic information did not affect physical activity of the participants ^17^.

### The change in risk factor levels are clinically meaningful

Finally, the change in health behavior observed among individuals at high ASCVD-risk in the current study did translate into significant improvement of blood lipid and blood pressure profiles. The majority of individuals who had weight loss during follow-up had not visited a physician. Nonetheless, their reduction in blood pressure was similar to the people who had consulted a physician during follow-up, i.e. 7 mmHg. Moreover, weight loss was associated with significant improvement of HDL-cholesterol and a 0.3 mmol/l reduction of serum triglycerides. While weight loss did not significantly affect LDL-cholesterol, majority of individuals who had consulted a physician had either been recommended further cholesterol tests or prescribed cholesterol-lowering therapy. Thus, their LDL-cholesterol dropped by 0.5 mmol/l during follow-up. These changes in risk factor profiles can be compared with previous meta-analyses studies indicating that 20% reduction of cardiovascular disease events can be achieved by a 10 mmHg reduction in systolic blood pressure or a 1 mmol/l reduction of LDL-cholesterol ^18^. Based on previous observational studies indicating that favorable lifestyle factors may compensate for high polygenic risk and post hoc analyses of primary prevention statin trials, suggesting a greater benefit from statins if the CHD PRS is high, the improvement of the risk factor profile, achieved by the GeneRISK-participants, if sustained, can be expected to facilitate a clinically meaningful reduction of ASCVD risk.

### Limitations of the study

Even if the GeneRISK–study has large sample size, it has certain limitations. All participants were between 45 and 65 years of age. Thus, the study does not provide direct information on disease prevention in young adults. Moreover, the study persons were from Finland and of European descent, and therefore the results may not be representative of more diverse populations. Finally, although the overall participation rate in GeneRISK was >70%, the attendance bias at follow-up may have impacted on the outcomes, i.e. individuals who are healthier were more likely to participate in the follow-up. However, the GeneRISK-study population as a whole has a similar ASCVD-risk profile as the Finnish population in general ^19,20^.

### Brief Summary & Conclusions

As guidelines for cardiovascular disease prevention typically encourage actions based on combined absolute risk profiles rather than individual risk factors, utilizing polygenic risk information alongside clinical data provides a natural way to make the routinely used risk evaluation tools more precise, personalized and comprehensive. Our study demonstrates the power of a digital risk prediction tool facilitating the presentation of polygenic risk information to patients in a comprehensive way alongside clinical risk factors. Not only does combining genomic and clinical information provide a more precise estimate of the overall disease risk on an individual level, our study further shows that both types of risk data independently predict positive health behavior, i.e. the higher the risk, the more likely a positive change. Thus, adopting procedures and tools facilitating the use of both genomic and clinical data for ASCVD-prediction provides a new basis for enhanced next generation disease prevention.

## Data Availability

The GeneRISK data may be accessed through THL Biobank (https://thl.fi/en/web/thl-biobank).

## Contributions

EW and SamR were responsible for the conception and design of the study. PP, KH, and JP oversaw the recruitment and clinical characterization of study participants and data collection. JA was responsible for the coordination of the study and data management. NJ, SanR, IS, NM, PR, and JJP performed data analyses. All the remaining authors contributed to data interpretation. The first draft was written by EW and SamR. All authors reviewed, commented on, and approved the final version of the manuscript.

## Acknowledgements

The GeneRISK study was funded by Business Finland through the Personalized Diagnostics and Care program coordinated by SalWe Ltd (Grant No 3986/31/2013).

SamR was supported by the Academy of Finland Center of Excellence in Complex Disease Genetics (Grant No 312062), the Finnish Foundation for Cardiovascular Research, the Sigrid Juselius Foundation and University of Helsinki HiLIFE Fellow and Grand Challenge grants. JK was supported by the Academy of Finland Center of Excellence in Complex Disease Genetics (Grant No 312073), the Sigrid Juselius Foundation and University of Helsinki HiLIFE Fellow grant. JJP was supported by the Doctoral Programme in Population Health, University of Helsinki and The Finnish Medical Foundation. Further support for the study was received from the Finnish innovation fund SITRA (EW) and Finska Läkaresällskapet (EW). PR acknowledges support from the Doctoral Programme in Population Health, University of Helsinki.

## Competing interests

VS has received honoraria for consulting from Novo Nordisk and Sanofi. He also has ongoing research collaboration with Bayer Ltd. (All unrelated to the present study). PP holds stock in Abomix Oy. The remaining authors have no competing interests to declare.

## Central Illustration

**The GeneRISK-study in brief; study protocol and main results.**

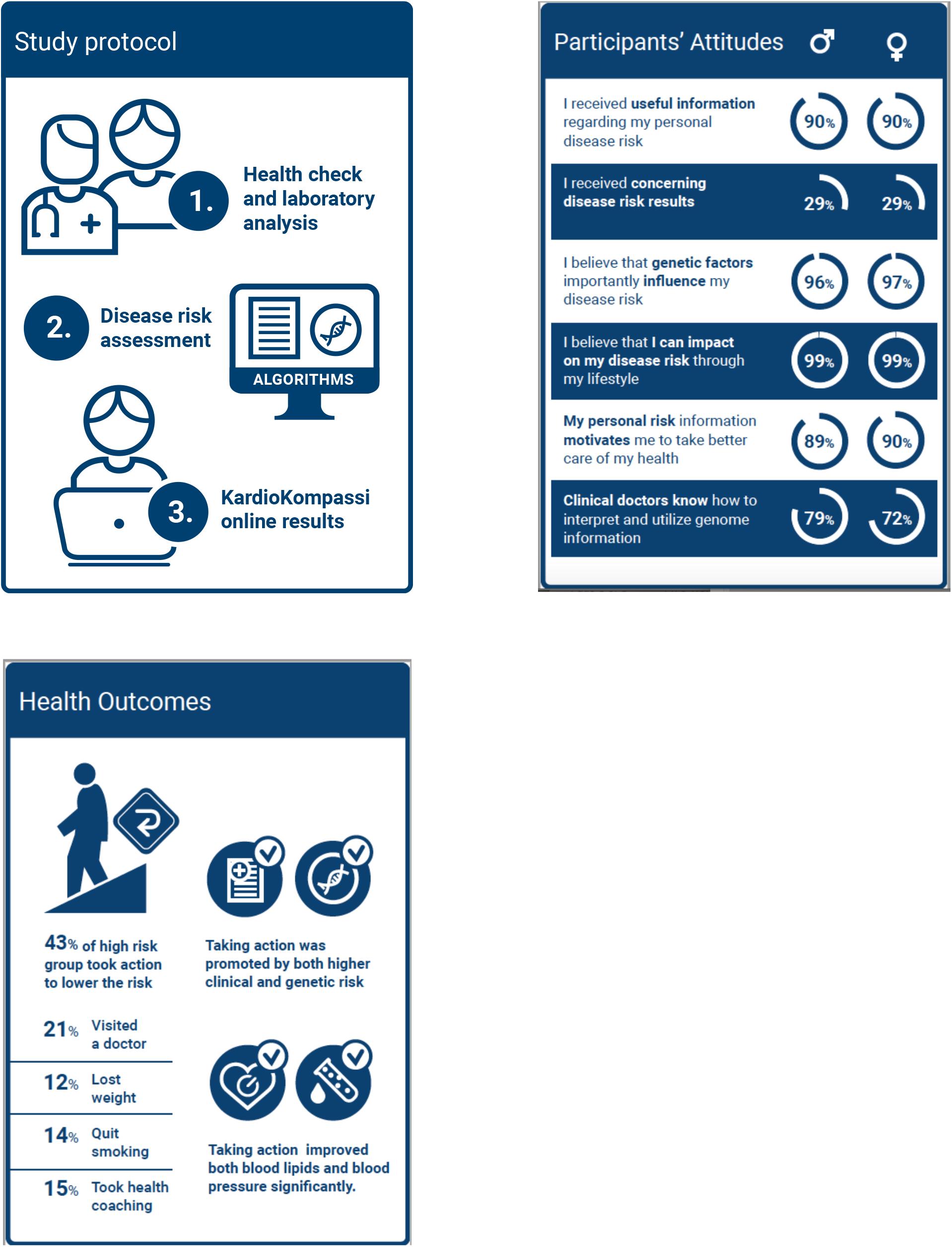

## Notes

### Author Declarations

The study protocol was approved by the Ethical Committee of the Helsinki and Uusimaa Hospital district.

